# The impact of paternal alcohol, tobacco, caffeine use and physical activity on offspring mental health: A systematic review and meta-analysis

**DOI:** 10.1101/2021.03.02.21252760

**Authors:** Kayleigh E Easey, Gemma C Sharp

## Abstract

**Background:** There is some evidence that paternal health behaviours during and around pregnancy could be associated with offspring health outcomes. However, the impact that paternal health behaviours during pregnancy can have on offspring mental health is understudied and remains unclear.

**Methods:** We conducted a systematic review and meta-analysis of articles in PubMed describing studies of potentially modifiable paternal health behaviours (tobacco smoking, alcohol consumption, caffeine consumption and physical activity) in the prenatal period in relation to offspring mental health.

**Results:** Ten studies were included and categorized by paternal health behaviour and offspring mental health outcome investigated. The narrative synthesis provided evidence of association between paternal health behaviours around pregnancy and offspring mental health problems, with the strongest evidence shown for tobacco use. Grouped by analysis type, two separate meta-analyses showed evidence of paternal smoking during pregnancy being associated with greater odds of ADHD in offspring (OR 1.42, 95% CI 1.02 to 1.99; HR 1.28, 95% CI 1.19 to 1.39).

**Conclusions:** Our review suggests that paternal tobacco smoking and alcohol consumption in the prenatal period are associated with poorer offspring mental health, particularly hyperactivity/ADHD. Future investigation using methods that allow stronger causal inference is needed to further investigate if these associations are causal.

## Introduction

The Developmental Origins of Health and Disease (DOHaD) literature has overwhelmingly focused on how maternal health behaviours during pregnancy may causally impact offspring health, including offspring mental health (1-3). Far less research has assessed the potential causal effect of paternal exposure (4, 5), although there is some evidence that paternal health behaviours during and around pregnancy could be associated with offspring health outcomes (6, 7).

Paternal traits and behaviours could influence offspring health through genetic inheritance and through environmental influences, both directly and indirectly (7). For example, germline transmission of epigenetic modifications could directly influence offspring health. However, there are also indirect pathways whereby paternal behaviours may influence the maternal environment, behaviour and physiology, which could then influence offspring outcomes. Direct prenatal paternal effects can only occur at pre-conception, whereas indirect effects can occur during pregnancy. Second-hand smoke exposure from paternal tobacco use during pregnancy is able and likely to impact the health of both mother and child. However, paternal use of alcohol, caffeine, or physical activity is unable to influence intrauterine development, except via a small indirect effect on maternal behaviour, such as by encouraging/discouraging exercise or not supporting mothers abstain from alcohol or caffeine during pregnancy. There is need to study these other paternal exposures during pregnancy to help to contextualise the maternal effect and as a negative control to explore causality. Negative control analyses are a method used to explore if associations are due to confounding or are likely to be causal (8), and are often the main reason why paternal exposures during pregnancy have been included in previous research (9, 10). A recent review distinguished between the impact of direct paternal biological and environmental effects, again highlighting the way paternal behaviour can impact offspring health (11). This review was broad, focusing on outcomes of male fertility, early pregnancy complications as well as fetal and postnatal outcomes. Although the previous review did find evidence of paternal factors having a detrimental effect on offspring neurodevelopmental disorders, this was shown only in relation to paternal exposures such as medication use or diagnoses of poor health. It was therefore not designed to specifically capture either mental health outcomes in offspring, or modifiable paternal health behaviours such as alcohol or tobacco use during pregnancy.

A recent review of the literature on paternal effects on child obesity and type 2 diabetes found some support for paternal influence on these outcomes, but also highlighted the paucity of high-quality research being conducted within this area (7). Another review from 2018 found evidence of associations of paternal age and paternal smoking with preterm birth, low birthweight and several congenital anomalies (12). The strongest associations shown were between paternal age and autism/autism spectrum disorders (pooled adjusted odds ratio per 5-year age increase: 1.25; 95% CI: 1.20 to 1.30) and schizophrenia (OR: 1.31; 95% CI: 1.23 to 1.38). However, it is unclear whether these and other psychiatric and mental health conditions are associated with potentially modifiable paternal health behaviours, like tobacco smoking, alcohol consumption, caffeine consumption and physical activity.

In this systematic review, we summarise the literature on associations of these paternal health behaviours in the prenatal period with offspring mental health outcomes. This provides insight into whether (and which) paternal prenatal health behaviours are likely to causally influence offspring mental health.

## Methods

This review was conducted according to PRISMA (Preferred Reporting Items for Systematic Reviews and Meta-analyses) guidelines (13), and preregistered on the Open Science Framework (*osf*.*io/adnbu*). We conducted a systematic search of PubMed to identify publications up until 01.03.2021.

### Search strategy

The search strategy included key words related to “paternal” (Paternal OR father*OR dad*OR partner*OR intergenerational) and “mental health” (‘mental health’ OR depress*OR anxiety OR mood OR internali?ing OR externali?ing OR conduct OR ADHD OR attention OR hyperactiv*OR ‘emotional problems’) and “offspring” (child*OR offspring OR son*OR daughter*) and “health behaviours” (smok*OR tobacco OR cigar*OR alcohol*OR caffeine OR coffee OR exercise*OR ‘physical activity’). We specified that these words should appear in the title or abstract of publications.

### Eligibility criteria

Studies were excluded if they were review articles with no original data or had not studied the exposures or outcomes of interest. Any source of mental health measure was included (e.g., self-report or parental report) and offspring outcomes could be measured at any age. Studies were only included if paternal exposures were measured at pre-conception or during pregnancy.

### Study selection and data extraction

One reviewer (KEE) assessed studies for inclusion/exclusion based on the title and abstract and full text. Another reviewer (GCS) assessed a random 10% of studies for inclusion/exclusion using the same criteria. For included studies, one reviewer (KEE) extracted data on study location, design, paternal exposure, exposure timepoint, mental health outcomes, offspring age, included covariates, sample size, whether both parents were studied, species studied, statistical methods and results. Where the above data was missing from manuscripts, we contacted authors of the original studies for further information. If included studies measured multiple mental health outcomes, the data were extracted separately for each outcome. We present the most covariate-adjusted results from the original study that are available.

### Meta-analysis

Fixed effects meta-analysis was conducted where appropriate (i.e., where exposures and outcomes were measured and defined similarly, and similar statistical tests were used to derive study estimates). Analyses were conducted using the *metan* command within Stata version 15. Between study heterogeneity was assessed using *I*^2^.

## Results

### Narrative synthesis

The initial search identified 963 articles, of which 943 were excluded based on title and abstract. Of the 20 that underwent full text review, 10 did not meet the inclusion criteria (did not measure paternal exposures in pregnancy/pre-conception). Of the ten included articles, eight studies were conducted in humans, and two in rodents. Offspring age varied from eight weeks to 24 years (four to 24 years in human studies only). Three studies were conducted in the USA, two in the UK, and one in each of Norway, Australia, Denmark, the Netherlands and Germany.

There was considerable heterogeneity between studies regarding measures of paternal exposure and offspring mental health outcome, length of follow-up, statistical methods used, and confounders adjusted for. Of the 10 included studies, seven included both maternal and paternal exposures using paternal exposures as a negative control analyses to explore causality (9, 14-19). The remaining three studies measured paternal exposure only (20-22).

**Figure 1.**
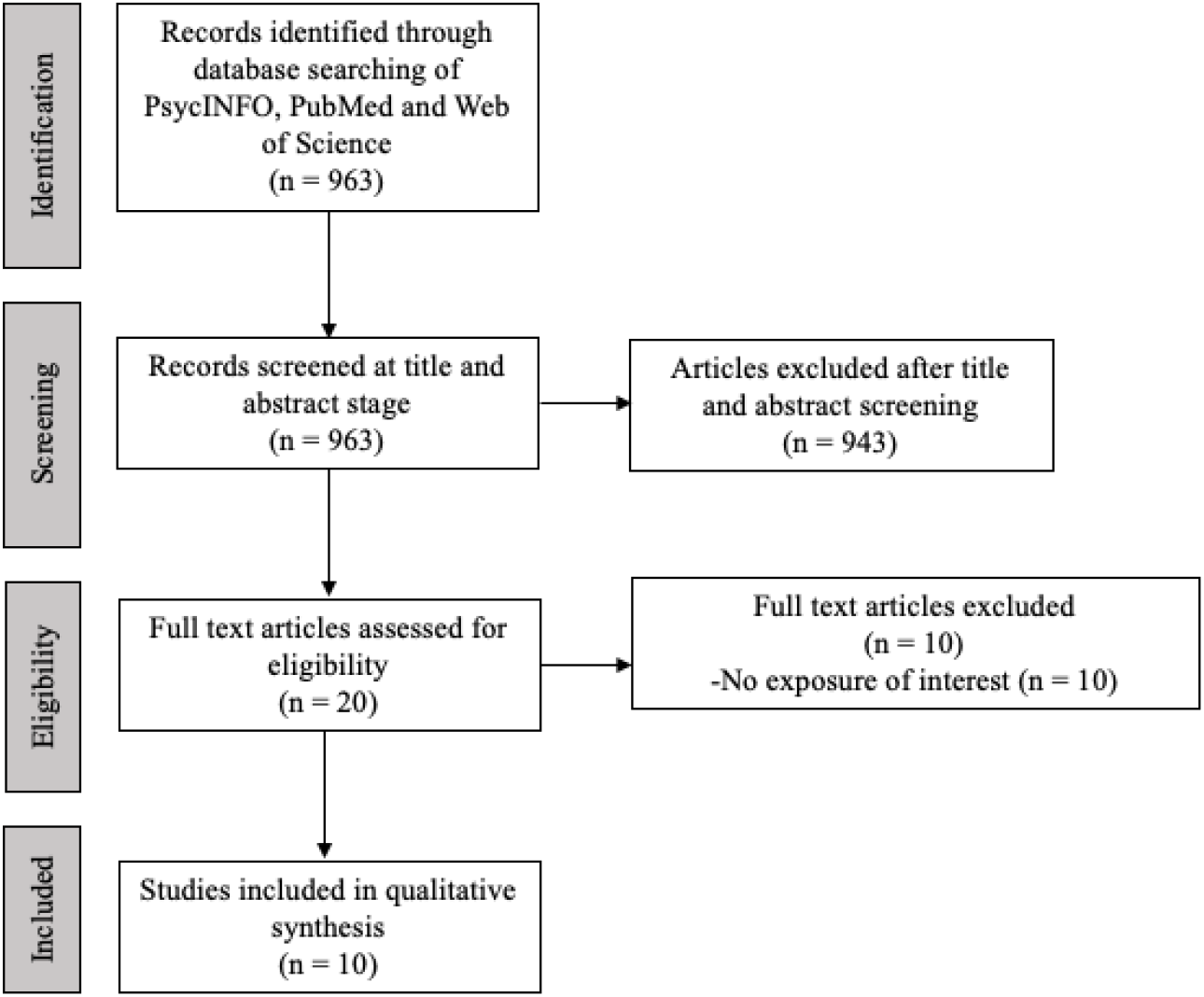
Flowchart of search strategy

**Figure 2.**
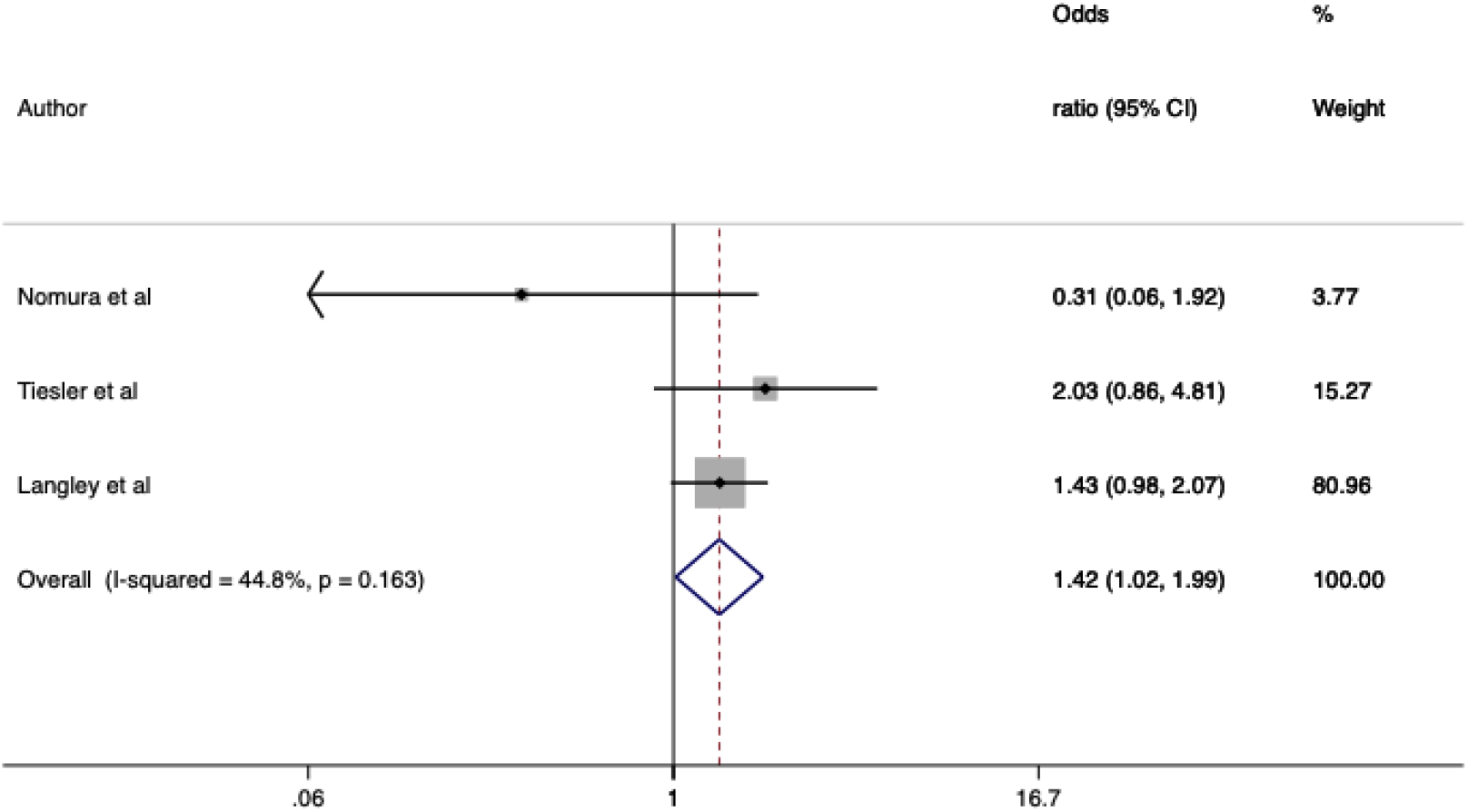
Forest plot of associations between paternal smoking in pregnancy and offspring ADHD of studies reporting odds ratios

**Figure 3.**
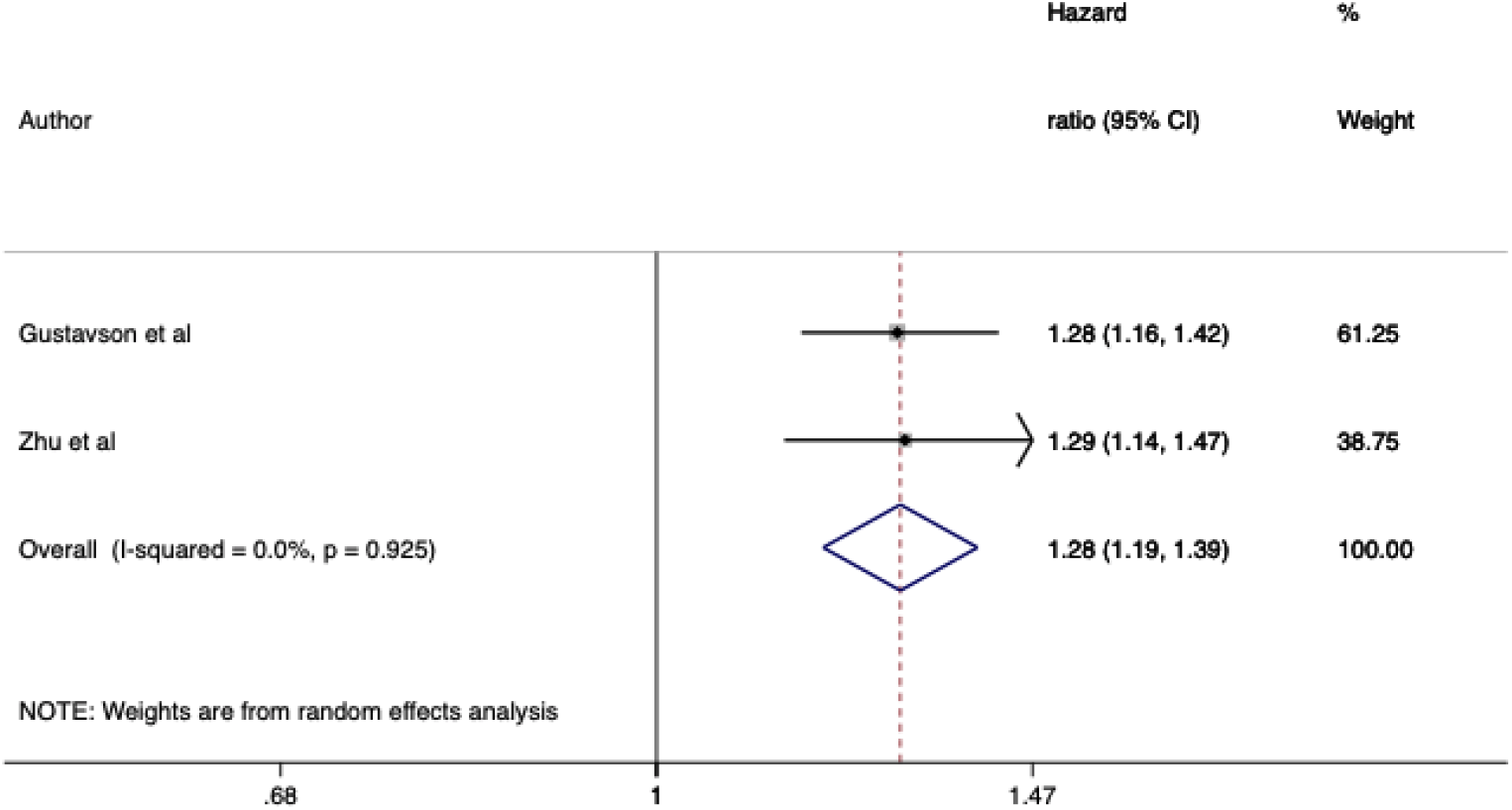
Forest plot of associations between paternal smoking in pregnancy and offspring ADHD of studies reporting hazard ratios

### Assessment tools used

Of the 10 included studies, seven studied paternal tobacco as an exposure, two studied alcohol, one studied physical activity, and there were no studies measuring caffeine. Seven studied paternal exposures during pregnancy and three just before pregnancy initiation. The studies considered seven different measures of offspring mental health (often more than one outcome was considered). ADHD and/or hyperactivity subscales were investigated by eight studies. Depression, anxiety, conduct disorder, emotional problems and a total problem score (combining multiple problems) were considered by one study each. Two studies investigated Oppositional Defiant Disorder (ODD). See table 1 for an overview of included full text studies.

**Table 1:**
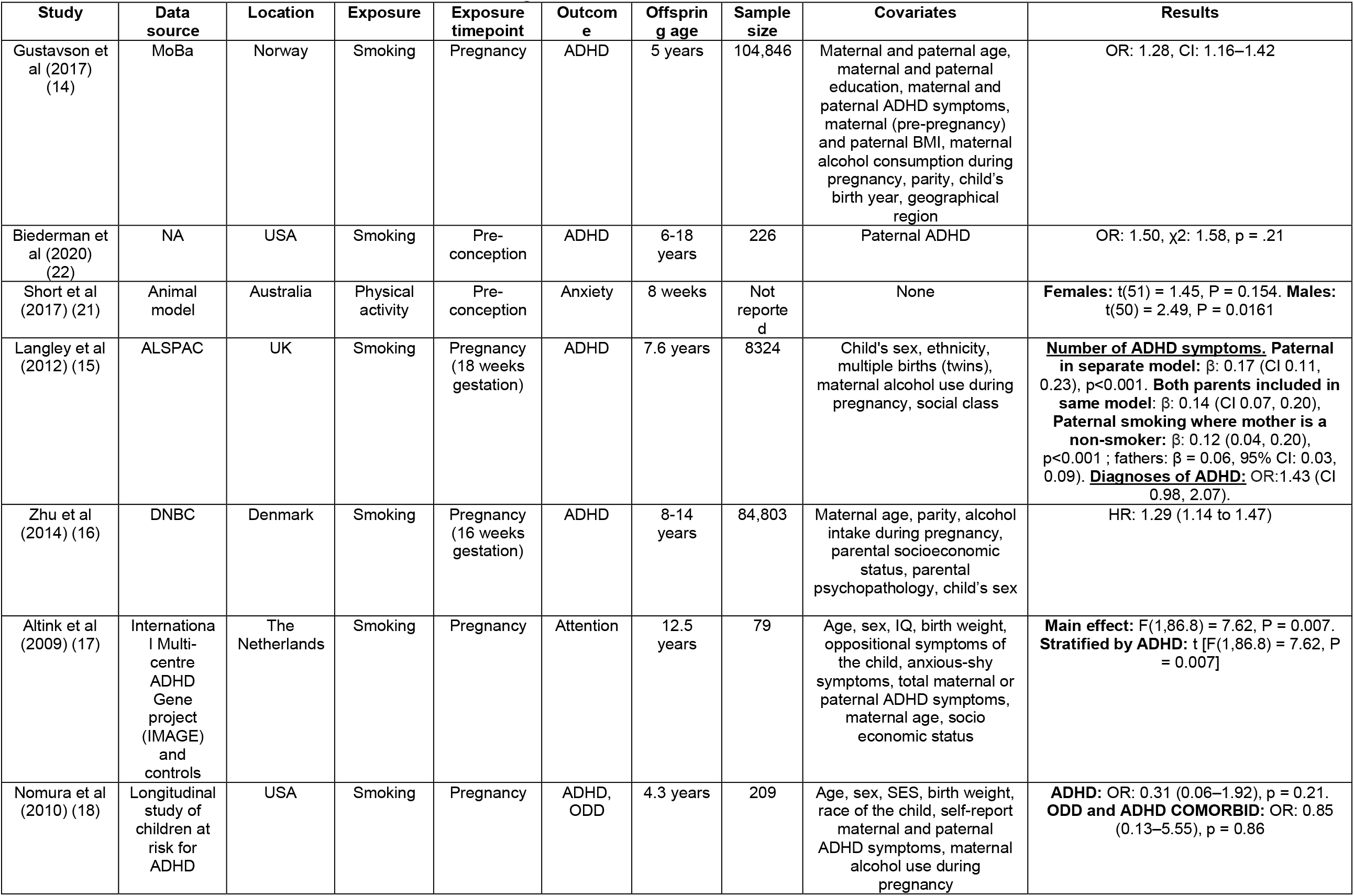
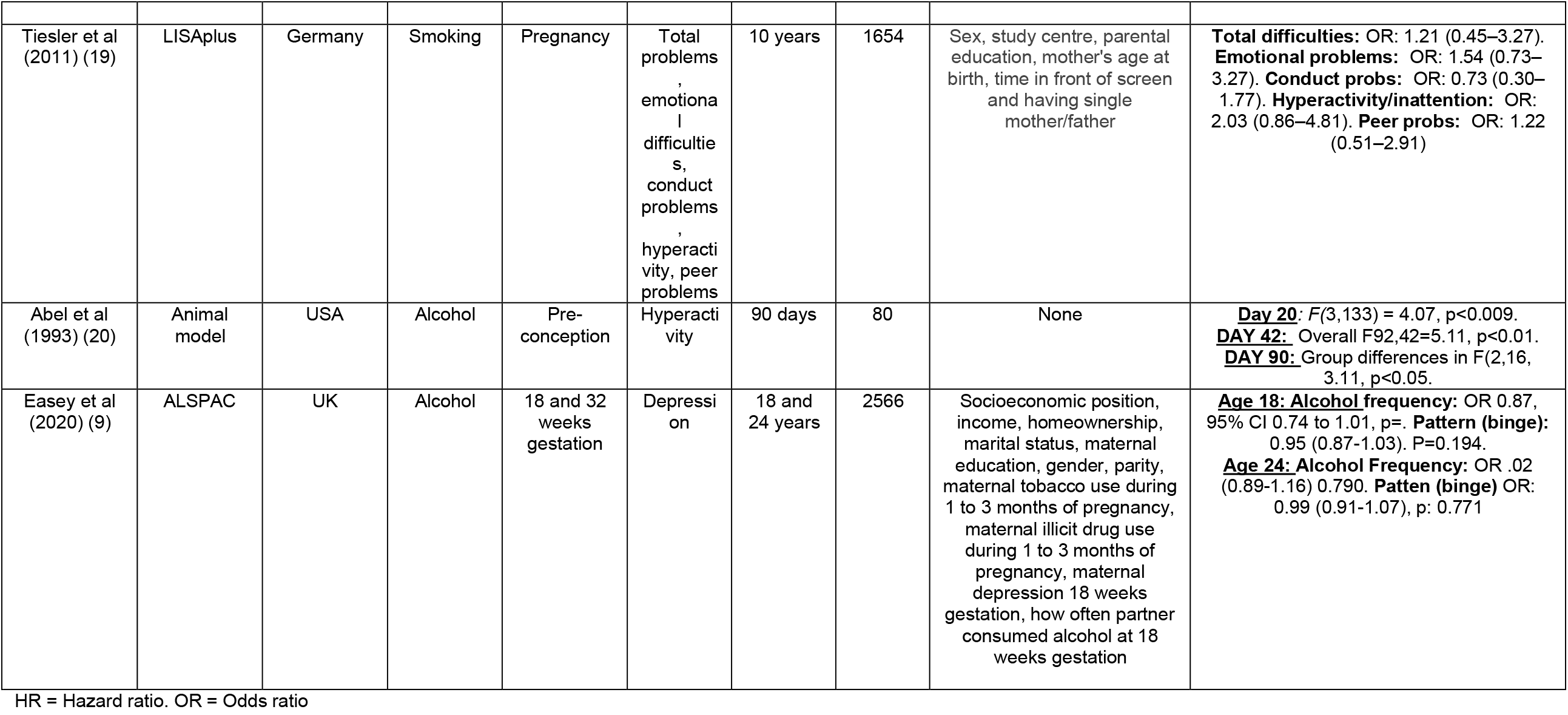
Studies included after full text screening

### Narrative synthesis

Of the 10 included studies, six (60%) found evidence of paternal health behaviours around pregnancy being associated with offspring mental health. Of these studies, five (50%) reported paternal substance use to be associated with increased offspring mental health problems; four of these studied paternal smoking, and the other studied alcohol. One study (10%) found increased amounts of paternal exercise to be associated with decreased anxiety in offspring. The four (40%) remaining studies reported no clear evidence of association; three of these studies considered paternal smoking and one considered paternal alcohol use.

### Meta-analysis

There was enough comparable data from studies of paternal smoking during pregnancy and offspring ADHD to conduct two meta-analyses: one of the 3 studies that reported odds ratios from logistic regression, and one of the 2 studies that reported hazard ratios. One additional study that met our criteria was not included, as following our request the corresponding author declined to provide additional statistical information required for the meta-analysis. Both meta-analyses found evidence that paternal smoking during pregnancy was associated with increased ADHD in offspring (logistic regression meta-analysis: OR 1.42, 95% CI 1.02 to 1.99, *I*^2^ = 45%; hazard models meta-analysis: HR 1.28, 95% CI 1.19 to 1.39, *I*^2^ = 0%).

## Discussion

The aim of this systematic review was to investigate the association between paternal health behaviours (alcohol, tobacco, caffeine, physical activity) around pregnancy and offspring mental health, and to quantify the degree to which published research has considered mental health in relation to these paternal prenatal health behaviours. Within our review, increased amounts of harmful substances of tobacco and alcohol during pregnancy were shown to be associated with detrimental offspring outcomes, however, increased physical activity in the pre-conception period was shown within one study to be protective for offspring mental health by reducing anxiety using a rodent model.

The majority of studies focused on paternal smoking above other paternal behaviours, and hyperactivity/ADHD was by far the most studied offspring outcome. Only three studies did not investigate tobacco use, and only two did not study hyperactivity/ADHD. This is unsurprising given that of the exposures we measured, paternal smoking is the only behaviour that can have an intrauterine effect (via passive smoke). This trend for research to focus on behavioural problems may not actually be unique to paternal exposures. A previous review of maternal alcohol use during pregnancy and offspring mental health has shown similar trends, with the majority of studies shown to measure offspring behavioural problems of conduct disorder, with less focusing on internalising problems (1). Of the studies included in the current review, half found paternal behaviours of smoking or alcohol use to be associated with negative offspring mental health outcomes, one found increased paternal physical activity to be potentially protective (associated with lower offspring anxiety) and the remaining four studies showed no clear evidence of association between paternal behaviours and offspring mental health. Paternal smoking during pregnancy was shown within the narrative synthesis and meta-analysis to be associated with increased rates off ADHD in offspring. There were no studies which evaluated paternal caffeine use in pregnancy, highlighting an under researched area in which future research is needed, to investigate if there are any (albeit potentially small) indirect effects of caffeine use on offspring mental health.

In general, our review suggests that paternal health behaviours around pregnancy may have an impact on offspring mental health, however, there are limitations within the included studies which can make causal interpretation challenging. Included studies varied wildly in terms of sample sizes, offspring ages, exposures and outcome measures. Furthermore, although we have presented results from fully adjusted models from each original study, there were inconsistencies in the confounders included in statistical models, which could contribute to inter-study heterogeneity in estimates. It is therefore unclear if residual confounding may be accounting for some results. Additionally, across included studies only three studies adjusted for the measured paternal health behaviour in the mother (e.g. studies of paternal smoking adjusting for maternal smoking). When studying any parental effect, mutually adjusted models help to account for exposure related assortative mating. Failing to make mutual adjustments for maternal and paternal exposures can result in bias from assortative mating (23, 24). By not adjusting for maternal pregnancy exposures, it cannot be certain that any associations with paternal exposures are not due to maternal contribution. Of the three studies that accounted for assortative mating through mutual adjustment, two showed paternal smoking to be associated with offspring ADHD, and the remaining study which measured paternal alcohol use and offspring depression showed no association. Future research using mutually adjusted models is needed to ascertain if the difference in associations shown between these behaviours are due to behaviour type and the inability for paternal alcohol use having a direct intrauterine affect, compared to passive smoke exposure as previously discussed.

Aside from the low number of studies found within this area, there are other limitations which must be considered when interpreting these results. Firstly, many of the included studies are observational and whilst they can identify associations, they do not alone provide evidence of causality. Causal inference is challenging due to the well-described problems of confounding and residual confounding in observational research (8). Secondly, which timepoint paternal health behaviours were measured within pregnancy/pre-conception was varied, with some studies only stating measures obtained for *during* pregnancy. This means we are unable to conclude if paternal exposure during any particular stage of pregnancy or pre-conception has a greater impact on offspring mental health. Lastly, in an attempt to assure the quality of included studies, articles were only included in this review if they were already published in peer-reviewed journals. However, the peer-reviewed scientific literature can suffer from publication bias whereby studies reporting null results are less likely to be published.

This review highlights the paucity of research that has investigated the association between paternal modifiable health behaviours around pregnancy and offspring mental health. Despite including four health behaviours as potential paternal exposures and using a broad definition of offspring mental health across any age, only ten studies were eligible for inclusion. By contrast, two reviews of maternal alcohol use and offspring mental health identified 26 (1) and 8 studies (25). The lack of research studying paternal effects and offspring mental health makes it challenging to draw conclusions from the available data.

In summary, we have identified 10 eligible studies of paternal health behaviours in relation to offspring mental health, six of which found some statistical evidence of association. Mirroring the focus in the literature on maternal effects on offspring mental health, most studies were of tobacco use during pregnancy, and most considered offspring ADHD/hyperactivity over internalizing behaviours. It is notable that we identified only a small number of studies that considered paternal exposures. This further highlights the imbalance of DOHaD research towards studies of maternal pregnancy exposures, which has been illustrated by other recent studies (4, 5). This review adds to research demonstrating the influence of paternal health and lifestyle on offspring health (6, 7, 11), and specifically the detrimental impact paternal behaviours around pregnancy may have on offspring mental health.

Given the potential for studies of paternal exposures to reveal important causal paternal effects, and to help contextualise the large body of maternal effects literature, further research is needed to investigate the causal impact of paternal health behaviours on mental health. Lack of evidence on the causal impact of paternal prenatal behaviours on offspring health may mean we are missing out on a potential pathway to reduce harm. Further research within this area could therefore influence paternal health warnings and advice for fathers during pregnancy and potential fathers to be during pre-conception.

## Data Availability

Data available from original studies.

## Acknowledgments

KEE and GCS work within the MRC Integrative Epidemiology Unit at the University of Bristol which is supported by the Medical Research Council (MRC) and the University of Bristol.

## Ethics approval and consent to participate

Not applicable.

## Consent for publication

Not applicable.

## Availability of data and materials

Not applicable.

## Authors’ contributions

The concept of this review was conceived by KEE and GCS. The protocol was written by, and screening and data extraction were performed by KEE and GCS. All authors read and approved the final manuscript.

### Abbreviations

DOHaD: The Developmental Origins of Health and Disease
ADHD: Attention Deficit Hyperactivity Disorder

## References

1. Easey KE, Dyer ML, Timpson NJ, Munafò MR. Prenatal alcohol exposure and offspring mental health: A systematic review. Drug Alcohol Depend. 2019;197:344–53.

2. Mamluk L, Jones T, Ijaz S, Edwards HB, Savovi ćJ, Leach V, et al. Evidence of detrimental effects of prenatal alcohol exposure on offspring birthweight and neurodevelopment from a systematic review of quasi-experimental studies. Int J Epidemiol. 2020.

3. Easey KE, Wootton RE, Sallis HM, Haan E, Schellhas L, Munafò MR, et al. Characterization of alcohol polygenic risk scores in the context of mental health outcomes: Within-individual and intergenerational analyses in the Avon Longitudinal Study of Parents and Children. Drug and Alcohol Dependence. 2021;221.

4. Sharp GC, Lawlor DA, Richardson SS. It’s the mother!: How assumptions about the causal primacy of maternal effects influence research on the developmental origins of health and disease. Soc Sci Med. 2018;213:20–7.

5. Sharp GC, Schellhas L, Richardson SS, Lawlor DA. Time to cut the cord: recognizing and addressing the imbalance of DOHaD research towards the study of maternal pregnancy exposures. J Dev Orig Health Dis. 2019:1–4.

6. Braun JM, Messerlian C, Hauser R. Fathers Matter: Why It’s Time to Consider the Impact of Paternal Environmental Exposures on Children’s Health. Curr Epidemiol Rep. 2017;4(1):46–55.

7. Sharp GC, Lawlor DA. Paternal impact on the life course development of obesity and type 2 diabetes in the offspring. Diabetologia. 2019;62(10):1802–10.

8. Gage SH, Munafò MR, Davey Smith G. Causal Inference in Developmental Origins of Health and Disease (DOHaD) Research. Annu Rev Psychol. 2016;67:567–85.

9. Easey KE, Timpson NJ, Munafò MR. Association of Prenatal Alcohol Exposure and Offspring Depression: A Negative Control Analysis of Maternal and Partner Consumption. Alcohol Clin Exp Res. 2020;44(5):1132–40.

10. Taylor AE, Carslake D, de Mola CL, Rydell M, Nilsen TIL, Bjørngaard JH, et al. Maternal Smoking in Pregnancy and Offspring Depression: a cross cohort and negative control study. Sci Rep. 2017;7(1):12579.

11. Montagnoli C, Ruggeri S, Cinelli G, Tozzi AE, Bovo C, Bortolus R, et al. Anything New About Paternal Contribution to Reproductive Outcomes? A Review of The Evidence. World J Mens Health. 2021.

12. Oldereid NB, Wennerholm UB, Pinborg A, Loft A, Laivuori H, Petzold M, et al. The effect of paternal factors on perinatal and paediatric outcomes: a systematic review and meta-analysis. Hum Reprod Update. 2018;24(3):320–89.

13. Moher D, Liberati A, Tetzlaff J, Altman DG, Group P. Preferred reporting items for systematic reviews and meta-analyses: the PRISMA statement. Ann Intern Med. 2009;151(4):264-9, W64.

14. Gustavson K, Ystrom E, Stoltenberg C, Susser E, Surén P, Magnus P, et al. Smoking in Pregnancy and Child ADHD. Pediatrics. 2017;139(2).

15. Langley K, Heron J, Smith GD, Thapar A. Maternal and Paternal Smoking During Pregnancy and Risk of ADHD Symptoms in Offspring: Testing for Intrauterine Effects. American Journal of Epidemiology. 2012;176(3):261–8.

16. Zhu JL, Olsen J, Liew Z, Li J, Niclasen J, Obel C. Parental smoking during pregnancy and ADHD in children: the Danish national birth cohort. Pediatrics. 2014;134(2):e382–8.

17. Altink ME, Slaats-Willemse DI, Rommelse NN, Buschgens CJ, Fliers EA, Arias-Vásquez A, et al. Effects of maternal and paternal smoking on attentional control in children with and without ADHD. Eur Child Adolesc Psychiatry. 2009;18(8):465–75.

18. Nomura Y, Marks DJ, Halperin JM. Prenatal exposure to maternal and paternal smoking on attention deficit hyperactivity disorders symptoms and diagnosis in offspring. J Nerv Ment Dis. 2010;198(9):672–8.

19. Tiesler CM, Chen CM, Sausenthaler S, Herbarth O, Lehmann I, Schaaf B, et al. Passive smoking and behavioural problems in children: results from the LISAplus prospective birth cohort study. Environ Res. 2011;111(8):1173–9.

20. Abel EL. Paternal alcohol exposure and hyperactivity in rat offspring: effects of amphetamine. Neurotoxicol Teratol. 1993;15(6):445–9.

21. Short AK, Yeshurun S, Powell R, Perreau VM, Fox A, Kim JH, et al. Exercise alters mouse sperm small noncoding RNAs and induces a transgenerational modification of male offspring conditioned fear and anxiety. Transl Psychiatry. 2017;7(5):e1114.

22. Biederman J, Fitzgerald M, Spencer TJ, Bhide PG, McCarthy DM, Woodworth KY, et al. Is Paternal Smoking at Conception a Risk for ADHD? A Controlled Study in Youth With and Without ADHD. J Atten Disord. 2020;24(11):1493–6.

23. Davey Smith G. Negative control exposures in epidemiologic studies. Epidemiology. 2012;23(2):350–1; author reply 1-2.

24. Lipsitch M, Tchetgen Tchetgen E, Cohen T. Negative controls: a tool for detecting confounding and bias in observational studies. Epidemiology. 2010;21(3):383–8.

25. Mamluk L, Edwards HB, Savović J, Leach V, Jones T, Moore THM, et al. Low alcohol consumption and pregnancy and childhood outcomes: time to change guidelines indicating apparently ‘safe’ levels of alcohol during pregnancy? A systematic review and meta-analyses. BMJ Open. 2017;7(7).

